# Early Crowdfunding Response to the COVID-19 Pandemic

**DOI:** 10.1101/2020.10.12.20211532

**Authors:** Sameh N. Saleh, Christoph U. Lehmann, Richard J. Medford

**Author notes:** Corresponding Author: Sameh N. Saleh, E.

## Abstract

**Background:** As the number of COVID-19 cases increased precipitously in the US, policymakers and health officials marshalled their pandemic responses. As the economic impacts multiplied, anecdotal reports noted the increased use of online crowdfunding to defray these costs. We examined the online crowdfunding response in the early stage of the COVID-19 pandemic in the US.

**Methods:** On May 16, 2020, we extracted all available data available on US campaigns created between January 1 and May 10, 2020 on GoFundMe and identified the subset of COVID-related campaigns using keywords relevant to the COVID-19 pandemic. We explored incidence of COVID-related campaigns by geography, by category, and over time and compared campaign characteristics to non-COVID-related campaigns after March 11 when the pandemic was declared.

**Results:** We found that there was a substantial increase in overall GoFundMe online crowdfunding campaigns in March, largely attributable to COVID-related campaigns. However, as the COVID-19 pandemic persisted and progressed, the number of campaigns per COVID-19 cases declined more than tenfold across all states. COVID-related campaigns raised more money, had a longer narrative description, and were more likely to be shared on Facebook than other campaigns in the study period.

**Conclusions:** Online crowdfunding appears to be a transient stopgap, predicated on the novelty of an emergency rather than the true sustained need of a community. Rather, crowdfunding activity is likely an early marker for communities in acute distress that could be used by governments and aid organizations to guide disaster relief and policy.

**Trial registration:** N/A

## Background

As the number of coronavirus 2019 cases (COVID-19) precipitously accelerated in March throughout the United States (US), government and public health departments marshalled their pandemic responses. Support efforts quickly became necessary as shortages of medical supplies and testing worsened by the continued spread of the virus affected health care organizations, medical offices, and nursing homes(1). While the first effects of the pandemic were health-related, the economic ramifications including rapidly increasing unemployment and loss of revenue quickly followed the spread of the disease and public health efforts to contain it(2,3). Temporary and permanent closure of small businesses, furloughing whole sectors of the economy (e.g. airline and restaurant industries), and food price inflation added social, financial, and medical strains to the population(4,5). Subsequent effects of the pandemic such as the increased number of deaths and associated funeral costs added additional financial stress to the lives of Americans.

Online crowdfunding has become an increasingly popular tool to finance medical treatment, respond to financial hardships and natural disasters, and defray the downstream economic impacts of personal healthcare costs(6,7). Despite anecdotal reports of the increased use of online crowdfunding to mitigate the economic impacts of COVID-19(8), details on if and how crowdfunding has been used for COVID-19 relief efforts remain unclear. We evaluate online crowdfunding campaigns in the US in the early stages of the COVID-19 pandemic on the GoFundMe platform, which represents 90% of the social crowdfunding market in the US as of 2018(9).

## Methods

We extracted (on May 16, 2020) all active US campaigns created between January 1 and May 10, 2020 on GoFundMe, the largest online crowdfunding platform in the US. The time frame reflects approximately 2 months before and after the World Health Organization (WHO) declaration of the pandemic on March 11, 2020. We used the BeautifulSoup library(10) to web scrape all data available for the GoFundMe campaigns and excluded all campaigns that originated outside the US. We list all extracted data fields in Appendix Table 1. We used keywords relevant to the COVID-19 pandemic to identify and label the subset of COVID-related campaigns (Appendix Table 2). The University of Texas Southwestern Human Research Protection Program Policies, Procedures, and Guidance did not require institutional review board approval as all data were publicly available.

We examined weekly new COVID-related campaigns juxtaposed with the weekly new COVID-19 cases both nationally and by state or territory. We evaluated the incidence of campaigns by state adjusted for new COVID-19 cases by week to determine the relationship between disease burden and campaign initiation. To allow for comparison among states, we defined week 0 as the first week that each state had at least 100 new weekly cases. We displayed all states, but only highlighted the top six states by overall campaigns by million. Given the presence of zip code level data, we also explored more granular community trends. We also evaluated changes in COVID-related campaigns per million for each state and campaign categories over time.

We compared campaign characteristics, including fundraising, campaign information, and campaign category, for COVID-19 and non-COVID-related campaigns over the same time period from March 11th to May 10th. We used Mann-Whitney U, X^2^, and Fisher’s exact testing where appropriate to determine significance. Alpha level of significance was set a priori at 0.05 and all hypothesis testing was two-sided. We did not adjust for multiple comparisons as this was an exploratory study and should be interpreted as hypothesis-generating. Analyses were performed using Python, version 3.7.2 (Python Software Foundation).

## Results

Of 310,695 total new campaigns created between January 1 and May 10, 2020, we analyzed 232,827 campaigns from the US across all categories. Twenty-two percent of these or 51,763 campaigns were identified as COVID-related and collectively raised $237,418,235 by May 10, 2020. The vast majority of COVID related campaigns were established after March 11th (50,828 or 98.2%). New GoFundMe campaigns had a peak in mid-March, mainly due to new campaigns related to or referencing COVID-19 and declined in April (Figure 1). This COVID-related increase in campaigns was observed before the national peak of new COVID-19 cases in April. Subsequently, the incidence of new campaigns declined while the incidence of new COVID-19 cases remained steady or declined slightly.

**Figure 1.**
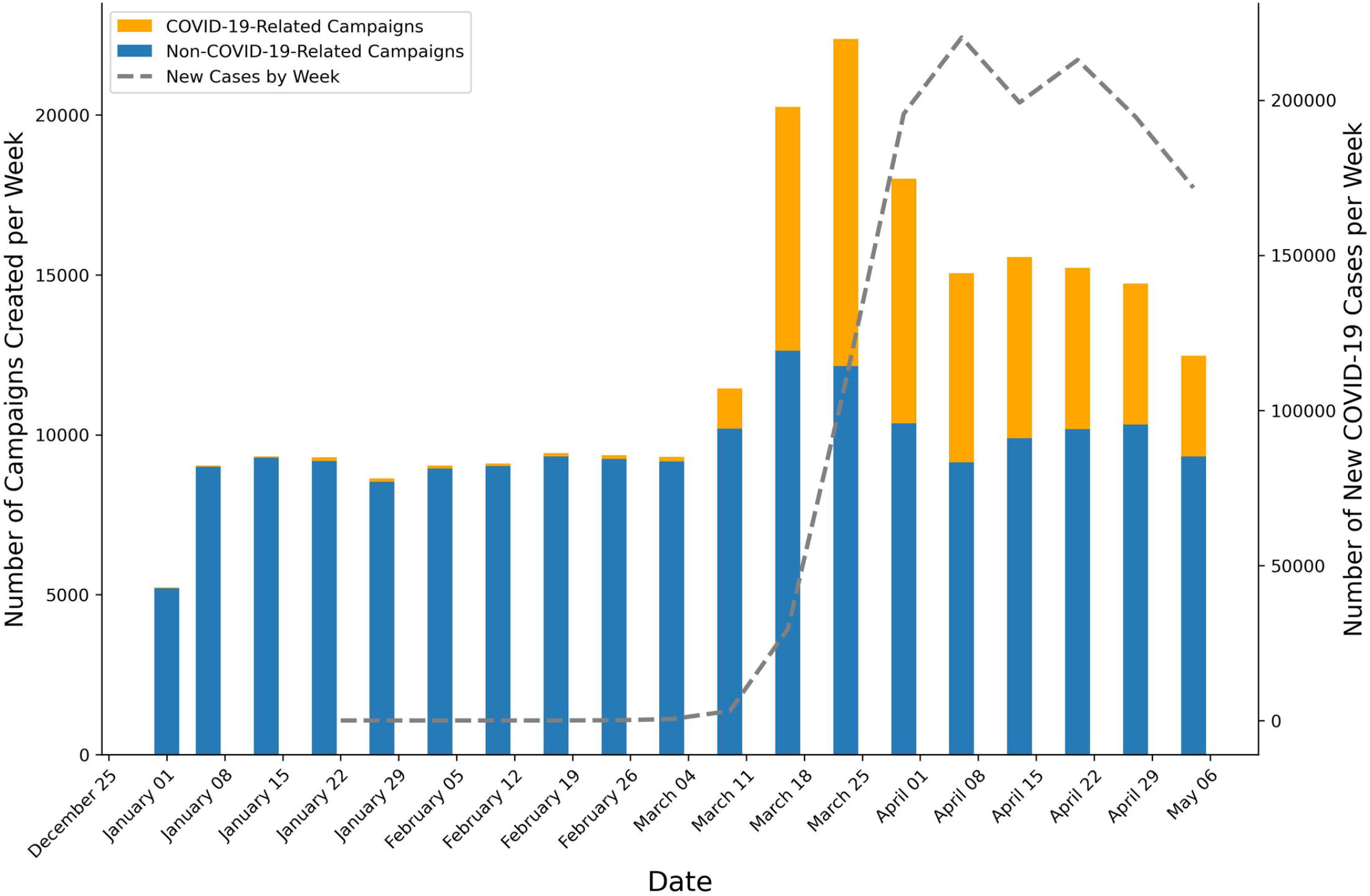
New campaigns by week versus incident COVID-19 cases by week. Yellow bars indicate COVID-related campaigns, blue bars indicate non-COVID-related campaigns, and the dotted line shows the number of incident cases.

Adjusting for the weekly incidence of COVID-19 cases, all US states had their peak of new COVID-related campaigns within 2 weeks of their first week when 100 new cases were diagnosed in that state. For many states, the rate of COVID-related campaigns per case dropped by at tenfold after the first 2 to 3 weeks (Figure 2). Population rich states that also had early spread of COVID-19 like New York, New Jersey, Massachusetts, Washington, and Illinois had the most COVID-related campaigns per million inhabitants (Figure 3), but were ultimately among the states with the lowest COVID-related campaigns per 1,000 cases during the study period.

**Figure 2.**
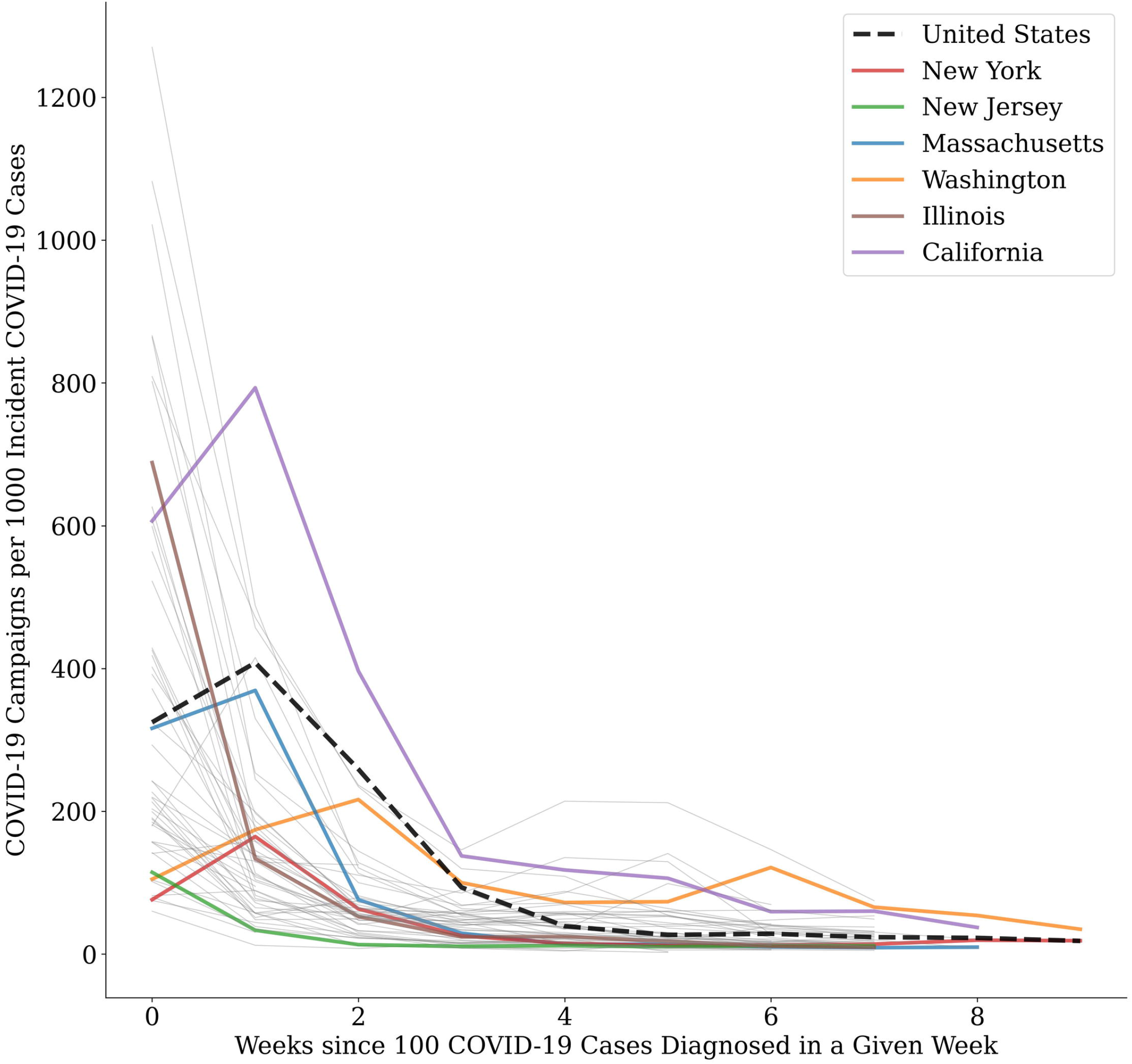
COVID-related campaigns per 1000 COVID-19 incident cases for each state. Week 0 is defined as the first week that a state had more than 100 cases to allow for direct comparison among states. All states are shown, but only the top 6 states for total COVID-19 cases per million are highlighted in color (as ordered from most to least in the legend).

**Figure 3.**
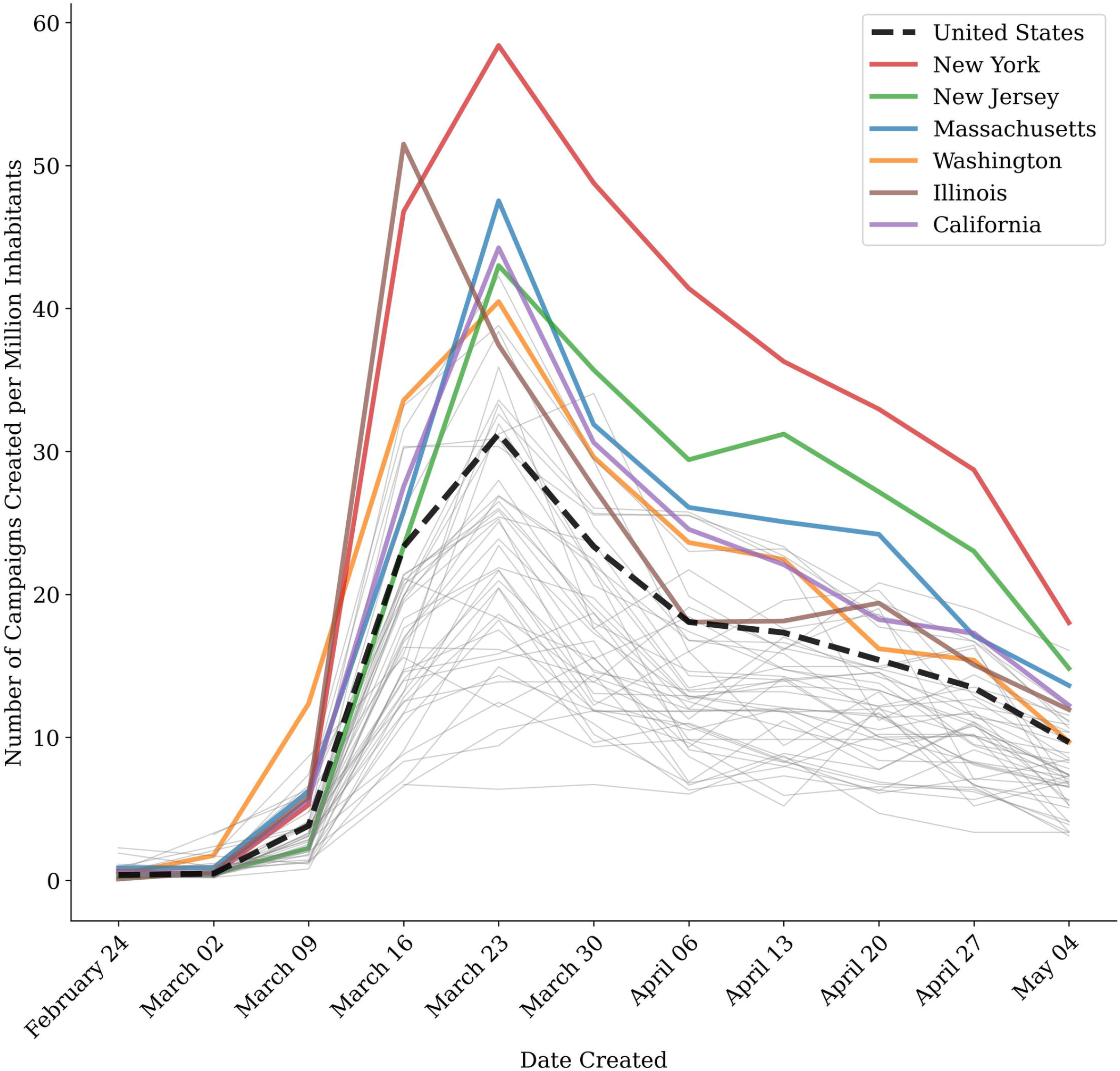
COVID-related campaigns per million by week by state. All states are shown, but only the top 6 states for total COVID-19 cases per million are highlighted in color (as ordered from most to least in the legend). Dates on the x-axis start from February 25th given how few COVID-related campaigns were present before this date.

From March 11th to May 10th 2020, non-COVID-related campaigns (n=91,631) raised a median of $625 ((interquartile range [IQR], $135–$2300) while COVID-related campaigns (n=50,828) raised a median of $930 IQR, $220–$3,075), nearly 50% more per campaign (p < 0.001). Fundraising goals were higher in COVID-related (median [IQR], $5,000 [$2,000 to $10,000]) than non-COVID-related (median [IQR], $4,000 [$1,250 to $10,000]) campaigns (p <0.001). Even with the higher fundraising goals, COVID-related campaigns (median [IQR], 25.0% [5.5% to 69.5%]) raised a higher percent of the fundraising goal than non-COVID-related campaigns (median [IQR], 22.2% [5.0% to 65.5%]) by date of data extraction (p <0.001). COVID-related campaigns (median [IQR], 23 [0–163]) had significantly more Facebook shares than non-COVID-related campaigns (0 [0–145]) (p <0.001). COVID-related campaigns also had a longer narrative in their campaign description, were more likely to be listed as a charity, and more likely to have the campaigner be the same as the beneficiary (Table 1).

**Table 1.** Baseline campaign characteristics stratified by COVID-related status. Only campaigns created after March 1, 2020 are included. Continuous variables are presented as median (IQR) and categorical variables as number (%).

|  | **Total Campaigns**<br>**n = 142,459** | **COVID-Related**<br>**Campaigns**<br>**n = 50,828** | **Non COVID-Related**<br>**Campaigns**<br>**n = 91,631** | **p<sup>a</sup>** |
| --- | --- | --- | --- | --- |
| **Fundraising** |  |  |  |  |
| Goal, median (IQR), \$ US | 5,000 (1,500 – 10,000) | 5,000 (2,000 – 10,000) | 4,000 (1,250 – 10,000) | <0.001 |
| Raised, median (IQR), \$ US | 722 (160 – 2,565) | 930 (220 – 3,075) | 625 (135 – 2,300) | <0.001 |
| Percent of goal funded, median (IQR) | 23.3 (5.2 – 66.9) | 25.0 (5.5 – 69.5) | 22.2 (5.0 – 65.5) | <0.001 |
| Donors, median (IQR) | 12 (3 – 35) | 14 (4 – 40) | 11 (3 – 32) | <0.001 |
| Amount per donation, median (IQR) \$US | 56.3 (35.4 – 86.8) | 59.9 (39.0 – 92.7) | 54.5 (33.8 – 83.5) | <0.001 |
| Met funding goal, No. (%) | 20,593 (14.5) | 7,302 (14.4) | 13,291 (14.5) | 0.48 |
| **Campaign Information** |  |  |  |  |
| Campaigner = beneficiary, No. (%) | 119,243 (83.7) | 43,445 (85.5) | 75,798 (82.7) | <0.001 |
| Is listed as a charity, No. (%) | 7,144 (5.0) | 3,629 (7.1) | 3,515 (3.8) | <0.001 |
| Narrative, median (IQR), words | 145 (84 – 240) | 187 (115 – 299) | 125 (72 – 207) | <0.001 |
| Narrative, median (IQR), characters | 56.3 (35.4 – 86.8) | 1,072 (652 – 1,729) | 691 (393 – 1,154) | <0.001 |
| Active donation days, median (IQR) | 23 (14 – 33) | 24 (15 – 38) | 22 (13 – 31) | <0.001 |
| Facebook shares, median (IQR) | 4 (0 – 152) | 23 (0 – 163) | 0 (0 – 145) | <0.001 |
| GoFundMe hearts, median (IQR) | 12 (3 – 33) | 14 (4 – 38) | 10 (3 – 31) | <0.001 |
| **Campaign Category (top 8 of 17)** |  |  |  |  |
| Accidents & Emergencies, No. (%) | 25,306 (17.8) | 10,705 (21.1) | 14,601 (15.9) | <0.001 |
| Medical Illness & Healing, No. (%) | 22,669 (15.9) | 8,484 (16.7) | 14,185 (15.5) | <0.001 |
| Funerals & Memorials, No. (%) | 19,577 (13.7) | 3,242 (6.4) | 16,335 (17.8) | <0.001 |
| Community & Neighbors, No. (%) | 14,237 (10.0) | 7,008 (13.8) | 9,027 (7.9) | <0.001 |
| Business & Entrepreneurs, No. (%) | 11,451 (8.0) | 6,458 (12.7) | 7,229 (5.4) | <0.001 |
| Animals & Pets, No. (%) | 11,158 (7.8) | 2,131 (4.2) | 9,027 (9.9) | <0.001 |
| Babies, Kids, & Family, No. (%) | 6,265 (4.4) | 1,990 (3.9) | 4,275 (4.7) | <0.001 |
| Volunteer & Service, No. (%) | 5,176 (3.6) | 2,643 (5.2) | 2,533 (2.8) | <0.001 |

<sup>a</sup> Given the non-Gaussian populations, we used Mann-Whitney U, Chi-squared, and Fischer's exact testing to detect statistical differences among the three groups.

Creators of COVID-related campaigns selected the following categories most commonly (in descending order by number of campaigns): 1) Accidents & Emergencies, 2) Medical Illness & Healing, 3) Community & Neighbors, 3) Business & Entrepreneurs, 5) Funerals & Memorials (Exhibit 5). Accidents & Emergencies as well as Community & Neighbors Medical Illness & Healing had the earliest peaks (Figure 4a). A week later, the category Business & Entrepreneurs peaked, but this appeared to be driven by an isolated spike on March 24th (Figure 4b). Medical Illness & Healing peaked next within the following 2 weeks. As these initial spikes decreased significantly, Funerals & Memorials campaigns lagged the first peak by 4 weeks. Compared to non-COVID-related campaigns after March 11th, COVID-related campaigns had more campaigns in Accidents & Emergencies, Community & Neighbors, and Business and Entrepreneurs and less campaigns in Funerals & Memorials and Animals & Pets.

**Figure 4.**
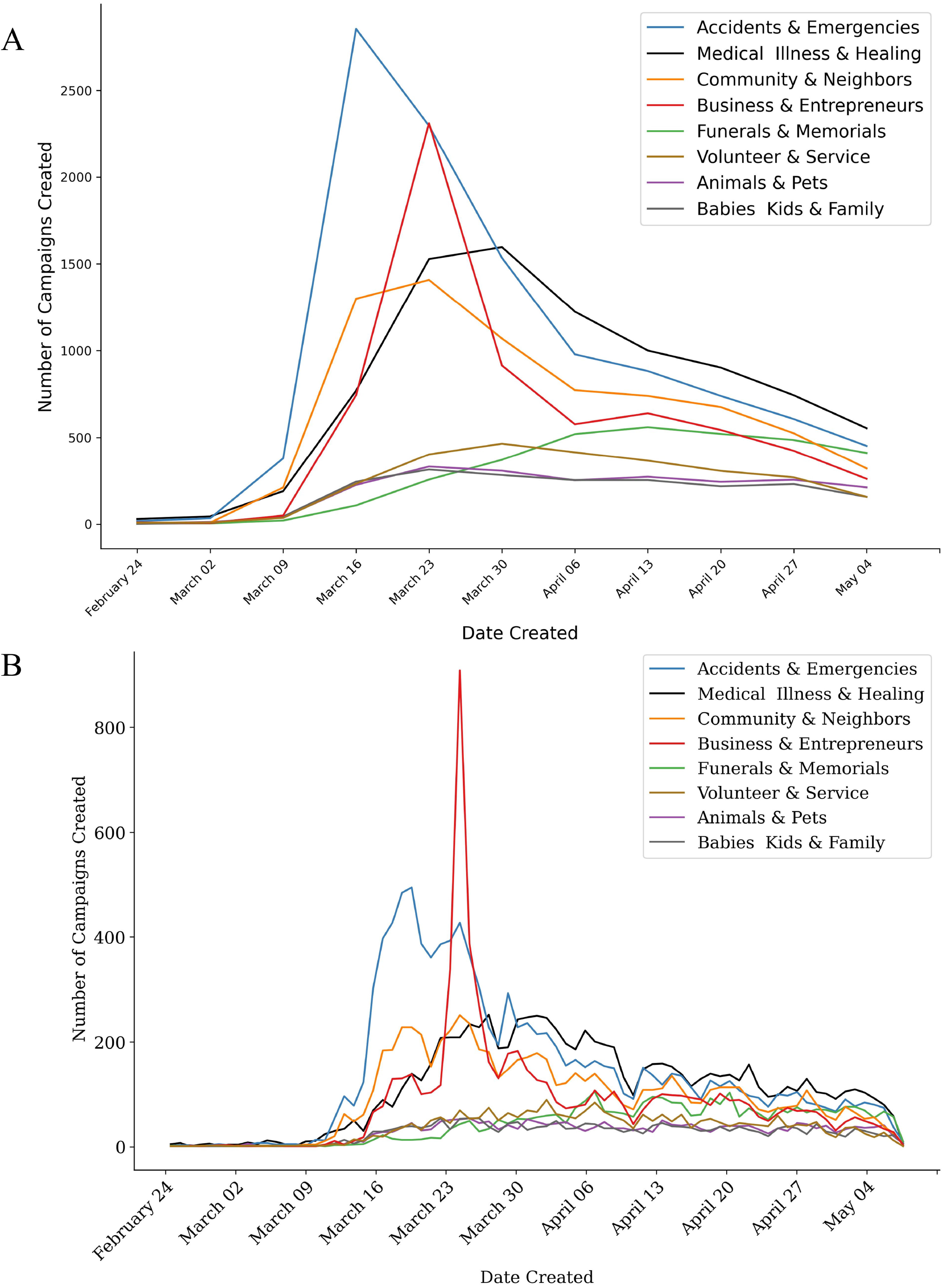
COVID-related campaigns per category a) by week and b) by day. Only the top 8 (of the 17 possible) categories are shown. The legend is ordered from most to least common category. Dates on the x-axis start from February 25th given how few COVID-related campaigns were present before this date.

## Discussion

Early in the COVID-19 pandemic, people were more likely to create COVID-19 campaigns. There was a substantial increase in overall GoFundMe online crowdfunding campaigns in March, mostly attributable to COVID-related campaigns. COVID-related campaigns raised more money than other campaigns, had a longer narrative description, and were more likely to be shared on Facebook. However, as the COVID-19 pandemic progressed, its novelty wore off. As COVID-19 became a familiar ubiquitous problem, the number of campaigns per COVID-19 cases declined across all states. This observation may be explained by potential campaign developers weighing the effort required to create a campaign with a shrinking potential for reward. Initially, the novelty and uniqueness of COVID-19 cases and with it, the chance of a campaign to “go viral”, reach many people, and generate large donations may have convinced potential campaign builders that COVID-related illness or financial hardships might be perceived as causes that others would support. As the issue became widespread and the ability to set a campaign apart as a worthy cause became more difficult, the effort (cost) of creating a campaign may have been outweighed by the perceived likelihood of funding. As others have found, the concept of worthiness and augmenting one’s “illness narrative” become important factors in generating campaign appeal, influence, and ultimately fundraising success(11–13). The states with the earliest disease burden had the least campaigns per case indicating a lack of a case-dependent response, supporting the hypothesis that potential campaign designers perceive common illnesses as less likely to receive the attention of donors over time.

While raising millions of dollars, our study suggests that online crowdfunding does not function as a true financial safety net. Medical expenses related to COVID-19 in the US are estimated to be in excess of $163 billion if 20% of the population were to be infected (on August 26, 2020, approximately 1.8% of the population had a confirmed infection)(14). Rather, crowdfunding more likely functions as a weathervane indicating a novel crisis. Further, crowdfunding is heavily swayed by marketing and storytelling as it points a majority of the funds towards a few cases rather than aiding the many(8,15).

The large number of categories for COVID-related GoFundMe campaigns tells a poignant story of the broad, destructive effect that COVID-19 had on society. Beyond creating the need to cover unexpected costly medical expenses, COVID-19 devastated small businesses, skyrocketed unemployment, created food and housing insecurities, generated unexpected expenses such as funeral costs, and increased debt(5). The campaign categories reflected the progression of the COVID-19 funding response with the earliest peaks for accidents and emergencies and the late growth and plateau for funerals and memorials. Spike in business campaigns was likely heavily based around the launch of the Small Business Relief Initiative on March 2(16).

As newly generated GoFundMe campaigns showed a temporal correlation to the pandemic’s effect on society, we contemplated that GoFundMe campaigns may actually serve as an early marker for any community in distress. To explore this hypothesis, we isolated campaigns from smaller communities that have been affected by a natural disaster including Puerto Rico (earthquake from January 6-7, 2020), Nashville (tornado from March 2-3, 2020), and Mississippi (tornado from April 12-13, 2020). We were able to discern an immediate peak for GoFundMe campaigns following each event (Appendix Figure 1).

While GoFundMe campaigns failed to meet the needs of those seeking support as demonstrated by the small fraction of funding goals met by data extraction date, it appears however to be an accurate, early marker for a community, region, state, or country in distress. This measure is not limited to medical emergencies and extends to natural disasters. Future research will show what type of negative events that affect communities may be reflected by a spike in GoFundMe campaigns. We propose the development of a measure using rapid rise in new GoFundMe campaigns as an early indicator of community distress.

While we were able to analyze the full available set of GoFundMe campaigns, which strengthened our study, it had several limitations. First, we used data from one crowdfunding platform only. While GoFundMe is the largest platform in the US by number of campaigns, our study may have suffered from selection bias. Because of the large number of campaigns, we are unable to validate that campaigns using COVID-related keywords were indeed designed to mitigate the effect of COVID-19. Moreover, while we note significant differences in campaign and funding characteristics between COVID-related and non-COVID-related campaigns, future work will be important to further understand reasons for these differences and their implications on long-term crowdfunding. Lastly, we only analyze the first few months of COVID-related campaigns; therefore, more longitudinal analysis will be important to understand more completely how peaks and troughs in COVID-19 incidence nationally and in particular, communities affect campaign frequency.

## Conclusions

Online crowdfunding activity leverages a galvanized public reaction early in a public health emergency like COVID-19. However, as disease spread persists and economic burden continues to grow nationally, the creation of new campaigns fades. Online crowdfunding appears to be a transient stopgap, predicated on the novelty of an emergency rather than the true sustained need of a community.

More importantly than its rather weak effect of mitigating the effect of the crisis, crowdfunding activity is likely an early marker for communities in acute distress. It could be monitored by local, state, and federal government agencies such as Federal Emergency Management Association (FEMA) as an early marker for emergencies and need for support. Future research will determine what emergencies generate the strongest signals in crowdfunding activities.

## Data Availability

The data that support the findings of this study are available upon request.

## Declarations

### Ethics approval and consent to participate

The University of Texas Southwestern Human Research Protection Program Policies, Procedures, and Guidance did not require institutional review board approval as all data were publicly available.

### Consent for publication

Not applicable

### Availability of data and materials

The data that support the findings of this study are available upon request.

### Authors’ contributions

Study concept and design: SS, RJM; Data acquisition: SNS; Analysis: SNS; Interpretation of data: SNS, CUL; Manuscript preparation: all authors. All authors read and approved the final manuscript.

## Acknowledgements

Not applicable

## References

1. Emanuel EJ, Persad G, Upshur R, Thome B, Parker M, Glickman A, et al. Fair Allocation of Scarce Medical Resources in the Time of Covid-19. N Engl J Med [Internet]. 2020 May 21 [cited 2020 Aug 24];382(21):2049–55. Available from: http://www.nejm.org/doi/10.1056/NEJMsb2005114

2. Almost 25 million jobs could be lost worldwide as a result of COVID-19, says ILO [Internet]. International Labour Organization; 2020 Mar [cited 2020 Aug 4]. Available from: https://www.ilo.org/global/about-the-ilo/newsroom/news/WCMS_738742/lang--en/index.htm

3. Tappe, Anneken. 30 million Americans have filed initial unemployment claims since mid-March. CNN Business [Internet]. 2020 Apr 30 [cited 2020 Jul 1]; Available from: www.cnn.com/2020/04/30/economy/unemployment-benefits-coronavirus/index.html;

4. Sherwan, Erik. Coronavirus wreaks havoc on Chinese food producers, sending inflation soaring. Fortune [Internet]. 2020 Feb 15 [cited 2020 Jul 13]; Available from: https://fortune.com/2020/02/15/china-coronavirus-business-impact-inflation-economy/

5. Nicola M, Alsafi Z, Sohrabi C, Kerwan A, Al-Jabir A, Iosifidis C, et al. The socio-economic implications of the coronavirus pandemic (COVID-19): A review. International Journal of Surgery [Internet]. 2020 Jun [cited 2020 Aug 24];78:185–93. Available from: https://linkinghub.elsevier.com/retrieve/pii/S1743919120303162

6. Saleh SN, Ajufo E, Lehmann CU, Medford RJ. Crowdfunding Medical Care: A Comparison of Online Medical Fundraising in Canada, the United Kingdom, and the United States. medRxiv [Internet]. 2020 Jan 1;2020.03.26.20044669. Available from: http://medrxiv.org/content/early/2020/03/30/2020.03.26.20044669.abstract

7. Bassani G, Marinelli N, Vismara S. Crowdfunding in healthcare. J Technol Transf [Internet]. 2019 Aug [cited 2019 Dec 24];44(4):1290–310. Available from: http://link.springer.com/10.1007/s10961-018-9663-7

8. Nathaniel Popper, Taylor Lorenz. GoFundMe Confronts Coronavirus Demand. The New York Times [Internet]. 2020 Mar 26 [cited 2020 Jul 13]; Available from: https://www.nytimes.com/2020/03/26/style/gofundme-coronavirus.html

9. Ainsley Harris. GoFundMe keeps gobbling up competitors, says it’s “very good for the market.” Fast Company [Internet]. 2018 Apr 4; Available from: https://www.fastcompany.com/40554199/gofundme-keeps-gobbling-up-competitors-says-its-very-good-for-the-market

10. BeautifulSoup [Internet]. Available from: https://www.crummy.com/software/BeautifulSoup/bs4/

11. Suzanne Woolley. American Health Care Tragedies Are Taking Over Crowdfunding. Bloomberg [Internet]. 2017 Jun 12; Available from: https://www.bloomberg.com/news/articles/2017-06-12/america-s-health-care-crisis-is-a-gold-mine-for-crowdfunding

12. Berliner LS, Kenworthy NJ. Producing a worthy illness: Personal crowdfunding amidst financial crisis. Social Science & Medicine [Internet]. 2017 Aug [cited 2019 Oct 20];187:233–42. Available from: https://linkinghub.elsevier.com/retrieve/pii/S0277953617300886

13. Lukk M, Schneiderhan E, Soares J. Worthy? Crowdfunding the Canadian Health Care and Education Sectors: Health Care and Education Crowdfunding. Canadian Review of Sociology/Revue canadienne de sociologie [Internet]. 2018 Aug [cited 2019 Oct 20];55(3):404–24. Available from: http://doi.wiley.com/10.1111/cars.12210

14. Bartsch SM, Ferguson MC, McKinnell JA, O’Shea KJ, Wedlock PT, Siegmund SS, et al. The Potential Health Care Costs And Resource Use Associated With COVID-19 In The United States: A simulation estimate of the direct medical costs and health care resource use associated with COVID-19 infections in the United States. Health Affairs [Internet]. 2020 Jun 1 [cited 2020 Sep 5];39(6):927–35. Available from: http://www.healthaffairs.org/doi/10.1377/hlthaff.2020.00426

15. Sumin Lee, Vili Lehdonvirta. New Digital Safety Net or Just More ‘Friendfunding’? Institutional Analysis of Medical Crowdfunding in the United States. SSRN. 2020 Mar 5;

16. GoFundMe Launches Relief Initiative to Help Small Business Impacted By COVID-19 in Partnership with Yelp. Business Wire [Internet]. 2020 Mar 24 [cited 2020 Jul 13]; Available from: https://www.businesswire.com/news/home/20200324005364/en/GoFundMe-Launches-Relief-Initiative-Small-Business-Impacted

