## Supplement for "Early Crowdfunding Response to the COVID-19 Pandemic"

**Appendix Table 1.** Data fields extracted for each campaign

| Url | Category | Category_IDs | Campaign_ID |
| --- | --- | --- | --- |
| Amount_Raised | Goal | Currency_Code | Number_of_Donors |
| Title | Text | Date_Created | Date_Launched |
| Date_of_Last_  Donation | GFM_Hearts | FB_Shares | Location |
| State | Country | Zip Code | is_Charity |
| Campaigner_SameAs_Beneficiary |  |  |  |

**Appendix Table 2.** Keywords to identify subset of COVID-related campaigns

| 2019-nCoV | corona | coronavirus |
| --- | --- | --- |
| COVID | COVID19 | COVID-19 |
| nCoV-2019 | SARSCoV2 | SARS-CoV-2 |
| pandemic | epidemic | outbreak |

**Appendix Figure 1.** Examples of incidence of new campaigns for different geographies with natural disasters.

A. Puerto Rico. Earthquake, January 6-7, 2020.

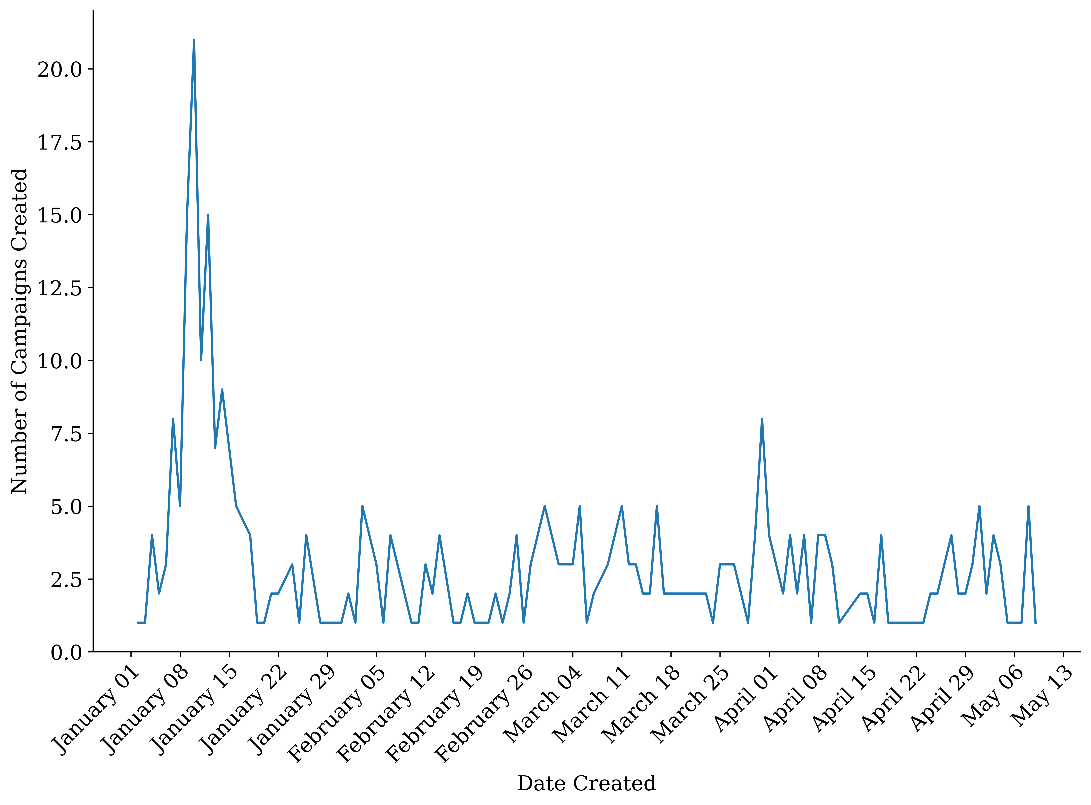

B. Nashville. Tornado from March 2-3, 2020.

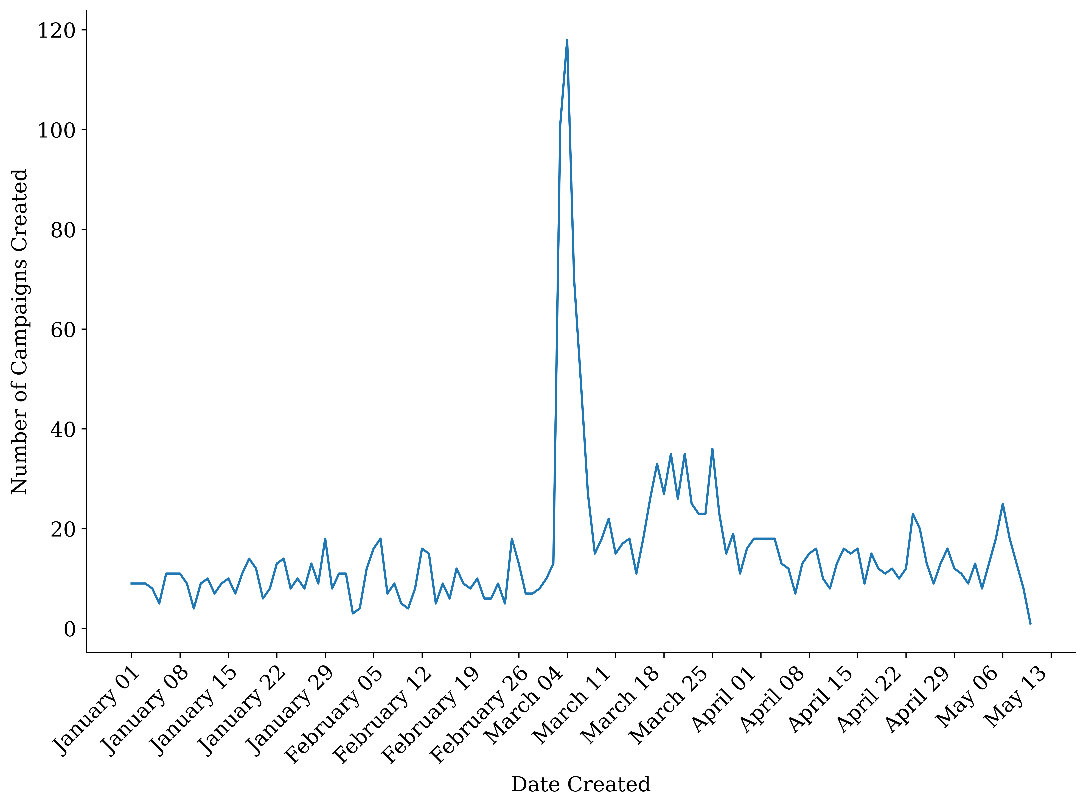

C. Mississippi. Tornado from April 12-13, 2020.

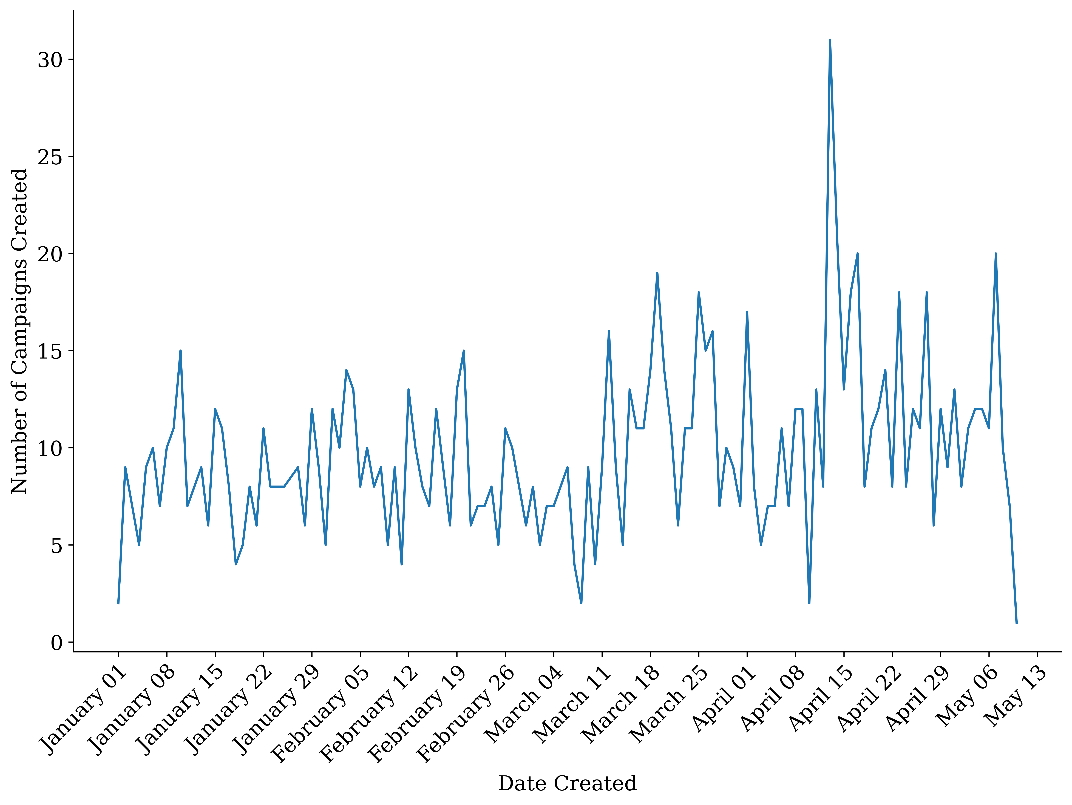
